# Factors Associated with Human Papillomavirus Vaccine Uptake Among Undergraduate Students in a Nigerian University: A Cross-Sectional Study

**DOI:** 10.1101/2025.10.15.25338122

**Authors:** Aisha Funmi Lawal, Amudalat Issa, Olayinka Rasheed Ibrahim, Adedapo M. Adesokan, Mohammed Jamiu Saka, Gordon Kayode Osagbemi

## Abstract

**Background:** Despite the proven effectiveness of human papillomavirus (HPV) vaccination in the prevention of HPV-related malignancies, vaccine uptake remains low in many low- and middle-income countries, including Nigeria. This study assessed the level of HPV vaccine uptake and associated factors among undergraduate students at a Nigerian university.

**Methods:** A descriptive cross-sectional study was conducted among 410 undergraduates selected through a multi-stage sampling technique between July and September 2023. Data were collected using a structured, self-administered questionnaire and analysed with SPSS version 26.

**Results:** Of the 410 male and female participants, 1.5% (6/410) received the HPV vaccine, all of whom were female (2.8% of the female respondents). Vaccine uptake was associated with knowing where to obtain the vaccine (p<0.001) and sex (p=0.037). Maternal levels of education, knowledge of the HPV vaccine, age, knowledge of HPV, and socioeconomic class were not related to vaccine uptake. Common reasons for non-uptake of the HPV vaccine included lack of awareness (63.2%; 211/334) and not knowing where to obtain the vaccine (19.2%; 64/334).

**Conclusion:** The uptake of the HPV vaccine was very low, with a major barrier to vaccination being a lack of awareness about the vaccine.

**Contribution:** This study provides a need to strengthened collaboration between university authorities and health authorities in raising awareness, improving accessibility, and enhancing the integration of HPV vaccination services within university settings in Nigeria.

## Introduction

Human papillomavirus (HPV) is a small, double-stranded DNA virus that infects the epithelial tissues and remains the most common sexually transmitted infection globally. Infection is particularly prevalent among sexually active women aged 24-45 years.[1,2] Among the more than 100 known HPV serotypes, high-risk types—especially HPV-16 and HPV-18—are responsible for nearly 70% of cervical cancer cases globally.[3–6] Cervical cancer is the most significant long-term consequence of HPV infection and ranks as the fourth most common cancer among women globally and the second most common female malignancy in Nigeria.[7] In Nigeria, approximately 12,075 new cervical cancer cases and 7,988 related deaths occur annually.[8,9]

Vaccination against HPV is a proven primary prevention strategy, achieving up to a 90% reduction in HPV-related cancers where it is implemented effectively.[1,10,11] The World Health Organization (WHO) recommends vaccination of girls aged 9-14 years as the primary target group, with males and females aged 15 years and above as secondary targets.[1] The vaccines are designed to prevent the serotypes commonly associated with the cancers, especially cervical cancer, and are most effective when administered before sexual exposure, as younger individuals mount a stronger immune response to the vaccine antigens.[12,13]

With the high burden of cervical cancer, the uptake of an effective preventive strategy such as HPV vaccination is expected to be high. In contrast, the vaccine uptake has persistently been low, especially in low- and middle-income countries, where HPV vaccines are yet to be free [11,13,14]. Despite its approval in Nigeria since 2009, HPV vaccine uptake remains low, with less than 10% reported in many parts of the country.[14–16] Uptake is usually confined to female adolescents in secondary schools, leaving a large segment of the at-risk population— particularly young adults in tertiary institutions—unreached. University students represent a critical secondary target for HPV vaccination due to their age, increasing sexual activity, and potential role as future advocates for preventive behaviour. However, data on HPV vaccine uptake and associated factors among this group are limited. This study, therefore, aimed to assess the level of HPV vaccine uptake and identify factors associated with uptake among male and female undergraduate students in a Nigerian university, prior to the integration of HPV vaccination into the national routine immunization schedule. Findings from this study are expected to provide valuable insights for policymakers and health authorities in strengthening HPV vaccination strategies and awareness campaigns among young adults in Nigerian tertiary institutions.

## Methods

### Study design and settings

This was a descriptive cross-sectional school-based study conducted from July to September 2023 among undergraduates at the University of Ilorin, Nigeria. The university, established in 1976, is located in Ilorin, Kwara State, North Central Nigeria. It has a health facility that provides outpatient and emergency services, excluding surgery, to students and staff of the university.

### Sample size determination

Fisher’s formula was used for estimating the minimum sample size required for the study (for minimum sample size calculation when the population is greater than 10,000)[17] Using a prevalence of 50% of the target population and a tolerable margin of error of 5% (0.05), a minimum sample size of 384 participants was obtained. However, 410 participants were finally recruited for the study.

### Sampling techniques

A multi-stage sampling technique was used to select the respondents for the study. In the first stage, six faculties—representing approximately one-third of the sixteen faculties in the university—were selected using simple random sampling by balloting. In the second stage, one or two departments were randomly selected from the chosen faculties, based on the proportion of departments within each faculty, resulting in a total of eight departments. The number of students recruited from each department was determined using a proportionate allocation based on the student population in each department. Within each selected department, students were further stratified by academic level (100, 200, 300, and 400 levels). In the third stage, systematic sampling was used to select eligible students from each stratum. The sampling interval was calculated by dividing the total number of students in each stratum by the required sample size as determined through proportionate allocation. The first respondent in each stratum was selected by simple random sampling through balloting, after which subsequent respondents were chosen systematically by adding the sampling interval until the desired number of participants was reached.

### Data collection

The research instrument used in this study was a structured, self-administered questionnaire developed following a comprehensive literature review. The questionnaire comprised the sociodemographic characteristics of the students, accessibility, and uptake of the HPV vaccine. The questionnaire also assessed students’ knowledge of HPV infection and reasons for non-uptake. For consistency, the questionnaire was checked for reliability and scored a Cronbach’s Alpha of 0.724, affirming its reliability. The questionnaires were pretested at a private University in the same town, about 25 kilometres away from the study site. This was done to identify problems with the instruments, and necessary modifications were made prior to the final administration.

### Data analysis

Data were computed and analysed using the Statistical Package for the Social Sciences (SPSS) version 26. The analysed data were presented in prose, frequency tables, and other appropriate charts. Univariate analysis was conducted to obtain descriptive statistics and frequencies, expressed as percentages. Bivariate analysis was used to test for associations between variables using cross-tabulations (chi-square). A confidence level of 95% was used, and a p-value of < 0.05 was considered significant.

### Ethical considerations

The University of Ilorin Ethical Review Committee granted ethical approval for this study (approval number UERC/ASN/2023/256). For participants under 18 years of age, written informed consent was secured from their parents in addition to their assent. For participants 18 years and above, a written informed consent was obtained for this study. The study protocol conformed to the ethical principles set forth in the 1975 Declaration of Helsinki. All collected data was handled with strict confidentiality.

## Results

### General characteristics of the study participants

The study comprised undergraduates aged 15 years and above, with a male-to-female ratio of 1:1.2. Most participants were 20 to 24 years of age (74.4%), unmarried, from the upper and middle socioeconomic classes, and were in the 100 to 300 level (**Table 1)**.

**Table 1:** General characteristics of the study population.

| Variable | Categories | Frequency (n=410) | Percent (%) |
| --- | --- | --- | --- |
| Age categories (years) | 15 to 19 | 73 | 17.8 |
|  | 20 to 24 | 305 | 74.4 |
|  | >25 | 32 | 7.8 |
| Gender | Male | 182 | 44.4 |
|  | Female | 228 | 55.6 |
| **Social class** | Upper | 302 | 73.7 |
|  | Middle | 89 | 21.7 |
|  | Lower | 19 | 4.6 |
| **Department** | Law | 39 | 9.5 |
|  | Accountancy | 76 | 18.5 |
|  | Medicine | 62 | 15.1 |
|  | Engineering | 50 | 12.2 |
|  | Nursing | 54 | 13.2 |
|  | Counselling Education | 52 | 12.7 |
|  | Human Kinetics | 51 | 12.4 |
|  | Political Science | 26 | 6.3 |
| **Marital status** | Single | 396 | 96.6 |
|  | Married | 14 | 3.4 |
| **Level** | 100 to 300 | 298 | 72.7 |
|  | 400 to 600 | 112 | 27.3 |
| **Religion** | Islam | 214 | 52.2 |
|  | Christianity | 194 | 47.3 |
|  | Others | 2 | 0.5 |

### Human papillomavirus vaccine uptake in study participants

Of the 410 respondents, six (1.5%) had received the HPV vaccine. Five of the six participants had two doses, and one had three doses.

### Factors that are associated with HPV vaccine uptake

Factors associated with the HPV vaccine include participants not knowing where to obtain the vaccine (p < 0.001) and sex (p = 0.036). Age, socioeconomic class, marital status, religion, mothers’ educational levels, and knowledge of the HPV vaccine were not associated with HPV vaccine uptake (**Table 2**).

**Table 2:** Factors Associated with HPV vaccine uptake.

| Variable | Categories | n: 410 | HPV Vaccine Uptake |  | P value |
| --- | --- | --- | --- | --- | --- |
| Sociodemographic factors |  | (%) | Yes: n=6 | No: n=404 |  |
| Age | 15 to 19 | 73 (17.8) | 0(0.0) | 73 (100.0) | 0.167* |
|  | 20 to 24 | 305 (74.4) | 6(2.0) | 299 (98.0) |  |
|  | >25 | 32 (7.8) | 0(0.0) | 32 (100.0) |  |
| Sex | Male | 228 (44.4) | 0(0.0) | 182 (100.0) | 0.036 |
|  | Female | 228 (55.6) | 6(2.6) | 222 (97.4) |  |
| Socioeconomic class | Upper | 302 (73.7) | 5 (1.7) | 297 (98.3) | 0.701* |
|  | Middle | 89 (21.7) | 1 (1.1) | 828(98.9) |  |
|  | Lower | 19 (4.6) | 0(0.0) | 19 (100.0) |  |
| Marital status | Single | 396 (96.6) | 6(1.5) | 390 (98.5) | 1.000# |
|  | Married | 14 (3.4) | 0(0) | 14 (100.0) |  |
| Religion | Islam | 214 (52.2) | 4 (1.9) | 210 (98.1) | 0.754* |
|  | Christianity | 194 (47.3) | 2 (1.0) | 192 (99.0) |  |
|  | Others | 2 (0.5) | 0(0.0) | 2(100.0) |  |
| Mothers' level of education | Graduate/HND | 222 (54.1) | 5 (2.3%) | 217 (97.7%) | 0.302* |
|  | OND/Health tech | 77 (18.8) | 0 (0.0%) | 77 (100.0%) |  |
|  | Secondary | 53 (12.9) | 1 (1.9%) | 52 (98.1%) |  |
|  | Primary | 43 (10.5) | 0 (0.0%) | 43(100.0%) |  |
|  | No formal education | 15 (3.7) | 0 (0.0%) | 15 (100.0%) |  |
| Vaccine-related factors |  |  |  |  |  |
| Know where to obtain HPV vaccine | Yes |  | 6 (10.3) | 52 (89.7) | <0.001# |
|  | No |  | 0(0.0) | 323(100.0) |  |
| Knowledge of HPV | Good | 23 (5.6) | 1 (4.3) | 22 (95.7) | 0.294# |
|  | Poor | 387 (94.4) | 5 (1.3) | 382 (98.7) |  |
| Knowledge of HPV vaccine | Good | 27 (6.6) | 2 (7.4) | 25 (92.6) | 0.053# |
|  | Poor | 383 (93.4) | 4 (1.0) | 379 (99.0) |  |

OND-Ordinary National Diploma; tech-technology; \*likelihood ratio; #-Fisher's Exact test.

## Reasons for non-uptake of the HPV vaccine by the study participants

Of the 334 respondents who provided information on reasons for non-uptake, the most common reasons cited were lack of awareness (63.2%), not knowing where to obtain the vaccine (19.2%), and a lack of trust in the vaccine’s efficacy (6.0%) (**Table 3**).

**Table 3:** Reasons for non-uptake of HPV vaccine by the study participants.

| Reasons for non-uptake of HPV vaccine (n=334) | Frequency | (%) |
| --- | --- | --- |
| Lack of awareness/information/recommendation | 211 | 63.2 |
| Do not know where to obtain the vaccine | 64 | 19.2 |
| Lack of trust in the efficacy of the vaccine | 20 | 6.0 |
| Vaccine is expensive | 8 | 2.4 |
| Vaccine not available in Nigeria | 12 | 3.6 |
| Fear of contracting HPV and other side effects concerns | 7 | 2.1 |
| No reasons | 12 | 3.6 |
| **Total** | **334** | **100.0** |

## Discussion

The HPV vaccine is one of the main preventive measures for cervical cancer. This study assessed the uptake of the HPV vaccine and explored associated factors of its uptake among undergraduates at a Nigerian university. Uptake of the HPV vaccine among the surveyed population is very low (1.5%), with only 2.8% of the females vaccinated. This finding is similar to what was obtained among secondary school students (1.9%) in the same geographical areas in 2015.[14] In Benin City, South-South Nigeria, only 1 out of 215 female adolescents (0.5%) attending secondary schools had received the HPV vaccine.[18] A comparative uptake level of 4% and 2.6% was reported in students in Lagos, Southwest Nigeria, and 4.1% among secondary school students in Ibadan in the same region.[19–21] The generally low uptake noted in this study is similar to that in many low- and middle-income countries, where the HPV vaccine is not yet available at the sub-national/national level. As of 2014, only 1.1% of girls aged 10-20 years in 84 low- and middle-income countries had received one or more doses of the HPV vaccine.[22] A similarly low uptake of 4.5% was reported in Pakistan when the vaccine had not been introduced at the national level.[23] All participants who received the HPV vaccine in this study were females, a similarity observed in many studies, as females are more likely to be vaccinated, even in countries where the vaccine is available to all.[9,24,25]. The higher uptake observed in females in this study may be attributed to the WHO’s emphasis on females as the primary target.

Five of the six subjects who received the HPV vaccine in the present study belonged to the high socioeconomic class, and one was from the middle socioeconomic class; their mothers all possessed tertiary education. This is not surprising, as vaccine acceptability has been reported to be associated with higher monthly income and higher levels of education in previous studies.[26,27] None of the participants from lower socioeconomic classes received the HPV vaccine. Lack of access to information on HPV infection and vaccines from parents, lack of financial accessibility, and their level of educational and social exposure may contribute to the non-uptake in this group of participants.

Most of those who had good knowledge of the HPV virus or the vaccine were, however, not vaccinated in this study. Good knowledge of a vaccine or its awareness may therefore not translate to high uptake when there are barriers to access and availability. There is a need for availability and improved access to the vaccine by all eligible individuals. This is a concern that the national/sub-national introduction of the vaccine may help to address, especially in Nigeria and other low-to medium-income countries where the highest burden of cervical cancer is.

Most of the participants in this study did not know where to obtain the vaccine, and this was associated with vaccine uptake. This may be due to a lack of awareness and sensitization of the vaccine and its availability. Lack of awareness, information, and recommendation, and not knowing where to obtain the vaccine were the major reasons for non-uptake of the HPV vaccine by participants in this study. Similarly, low uptake was observed among some adolescents in Lagos, Nigeria, which was attributed to a lack of awareness about vaccination centres and the high cost of the vaccine.

### Strengths and limitations

This is a recent cross-sectional analysis in a critical, under-researched population (undergraduate students), providing contemporary data on a national public health priority. However, this study has some limitations. Firstly, it is a cross-sectional study design, which cannot establish causality. Secondly, it is a single university, which limits its generalizability. Thirdly, the possibility of recall bias from the students’ responses. Finally, a qualitative study as part of this study may have provided a much broader perspective on the reasons for the uptake of the HPV vaccine.

## Conclusion

This study reveals that HPV vaccine uptake among undergraduate students in Ilorin, north-central Nigeria, is very low, mainly due to a lack of awareness and accessibility. We recommend the urgent strengthening of collaboration between university authorities and the Ministry of Health to improve awareness, accessibility, and integration of HPV vaccination services within university settings.

## Data Availability

The data associated with this study is available from the corresponding author upon a reasonable request

## Abbreviations

HPV: Human papillomavirus
WHO: World Health organization

## Declarations

### Ethics approval

Ethical approval was obtained from the University of Ilorin Ethics Review Committee and was conducted in accordance with the Declaration of Helsinki.

### Consent to participate

Informed consent and assent were obtained from all participants after a detailed study of information, as indicated in the method section.

### Consent for publication

All co-authors read the final version and provided their consent for publication.

### Authors’ contributions

Conceptualization: AFL, MJS,

Data analysis/interpretation: AFL, MJS, AI, ORI, GKO

Writing of Manuscript: AFL, MJS, AI, AA

Review the final version of the manuscript: AFL, MJS, AA, AI, ORI, GKO

